# Viability RT-PCR for SARS-CoV-2: a step forward to solve the infectivity quandary

**DOI:** 10.1101/2021.03.22.21253818

**Authors:** Enric Cuevas-Ferrando, Walter Randazzo, Alba Pérez-Cataluña, Irene Falcó, David Navarro, Sandra Martin-Latin, Azahara Díaz-Reolid, Inés Girón-Guzmán, Ana Allende, Gloria Sánchez

## Abstract

**Background:** Isolation, contact tracing and restrictions on social movement are being globally implemented to prevent and control onward spread of SARS-CoV-2, even though the infection risk modelled on RNA detection by RT-qPCR remains biased as viral shedding and infectivity are not discerned. Thus, we aimed to develop a rapid viability RT-qPCR procedure to infer SARS-CoV-2 infectivity in clinical specimens and environmental samples.

**Methods:** We screened monoazide dyes and platinum compounds as viability molecular markers on five SARS-CoV-2 RNA targets. A platinum chloride-based viability RT-qPCR was then optimized using genomic RNA, and inactivated SARS-CoV-2 particles inoculated in buffer, stool, and urine. Our results were finally validated in nasopharyngeal swabs from persons who tested positive for COVID-19 and in wastewater samples positive for SARS-CoV-2 RNA.

**Findings:** We established a rapid viability RT-qPCR that selectively detects potentially infectious SARS-CoV-2 particles in complex matrices. In particular, the confirmed positivity of nasopharyngeal swabs following the viability procedure suggests their potential infectivity, while the complete prevention of amplification in wastewater indicated either non-infectious particles or free RNA.

**Interpretation:** The viability RT-qPCR approach provides a more accurate ascertainment of the infectious viruses detection and it may complement analyses to foster risk-based investigations for the prevention and control of new or re-occurring outbreaks with a broad application spectrum.

**Fundings:** This work was supported by Spanish Scientific Research Council (CSIC), Generalitat Valenciana, and MICINN co-founded by AEI/FEDER, UE.

## Introduction

The rapid spread of severe acute respiratory syndrome coronavirus 2 (SARS-CoV-2) has led to an unprecedented global health and economic crisis. SARS-CoV-2 belongs to the *Coronaviridae* family, which includes enveloped RNA viruses causing respiratory, enteric, and systemic infections in a wide range of hosts, including humans and animals. Human coronaviruses have been traditionally considered responsible for endemic infections causing common cold symptoms, as in the cases of HKU1, 229E, OC43, and NL63 viruses, while more recently Middle East respiratory syndrome coronavirus (MERS-CoV) and SARS-CoV produced more severe epidemics in the Arabian Peninsula and in Asia. COVID-19 symptoms range from mild to severe, in which severe pneumonia and respiratory distress syndrome can lead to death. However, a significant number of infected people are asymptomatic, making the epidemiological control even more challenging.

SARS-CoV-2 is an airborne human pathogen primarily transmitted through droplets and aerosols, even though its detection in urine and faecal specimens raised the hypothesis of the possible fecal-oral transmission further sustained by the successful viral replication in cell culture.^(1,2)^ To control SARS-CoV-2 spread, extreme containment measures have been enforced worldwide along with several epidemiological surveillance strategies, which include tracing confirmed and suspected cases by clinical testing (e.g., SARS-CoV-2 nucleic acid or antigen tests on nasal or oral swabs or saliva samples), and monitoring community transmission by wastewater analysis (known as Wastewater Based Epidemiology, WBE).^(3)^

In this context, several molecular assays based on real-time reverse transcriptase polymerase chain reaction (RT-qPCR) have been developed to detect and quantify SARS-CoV-2 RNA in clinical and environmental samples. For instance, a test-based strategy (at least two consecutive negative RT-qPCR tests) has been widely adopted as a general public health guidance for release from (self-) isolation, reincorporation into the workplace, and patient transferral. However, COVID-19 patients can continue to shed viral RNA well beyond clinical recovery and persistent positive RT-qPCR does not necessarily indicate infectiousness.^(4)^ Besides being a rapid, easy-to-use, and cost-effective technique, RT-qPCR informs on the presence of viral RNA that does not correlate with infectivity, yet such testing is still being used as a surrogate marker of infectivity.^(5–8)^ On the contrary, viral replication in permissive cell line(s) represents the conclusive evidence to assess viral infectivity, and it has been readily available for SARS-CoV-2. Conversely, the facility requirements needed to handle SARS-CoV-2 infectious materials (biosafety level 3 laboratory, BSL-3), in addition to the low sensitivity and long turnaround time for results, typically from three to ten days, limited its extensive implementation for both clinical diagnosis and environmental risk assessment.^(9)^

Recently, novel molecular techniques, referred to as capsid integrity or viability qPCR assays incorporating viability markers such as monoazide dyes and metal compounds into qPCR-based methods, have been demonstrated to selectively remove false-positive qPCR signals deriving from free nucleic acids and virions with damaged capsids, finally allowing an estimation on viral infectivity.^(10)^ However, the application of such techniques for enveloped viruses has not fully elucidated as it has failed for avian influenza virus (IAV) and infectious laryngotracheitis virus (ILTV) while it has recently been optimized for porcine epidemic diarrhea coronavirus.^(11–13)^ Nonetheless, the implementation of this technique has not been explored for SARS-CoV-2.

## Methods

### Viral materials, viability markers and optimization of viability treatment

SARS-CoV-2 genomic RNA (VR-1986D™, ATCC, VA, US), gamma-irradiated (5 × 10^6^ RADs) (NRC-52287, BEI Resources, VA, US) and heat inactivated (65°C for 30 min) (NR-52286, BEI Resources, VA, US) viral particles preparations all obtained from isolate USA-WA1/2020 were used for initial screening of viability markers. Specifically, monoazide photoactivatable dyes and platinum compounds were initially screened as viability marker candidates using SARS-CoV-2 genomic RNA, gamma-inactivated, and heat inactivated SARS-CoV-2 suspensions. Viability marker stock solutions were prepared as follows and stored at −20 °C for later use: ethidium monoazide (EMA™, Geniul, Spain) was diluted in dimethylsulfoxide (DMSO) to 2·0 mM, PEMAX™ (Geniul, Spain) and propidium monoazide (PMAxx™, Biotium, CA, US) were diluted in nuclease-free water to 4·0 mM, platinum (IV) chloride (PtCl_4_; Acros Organics, NJ, US) and cis-diamineplatinum (II) dichloride (CDDP; Sigma-Aldrich, MO, US) salts were dissolved in DMSO to 1·0 M and further diluted in nuclease-free water to 50 mM. Viability assays were carried out by treating 300 µL of either genomic SARS-CoV-2 RNA (approx. 10^3^ gc/mL), gamma-inactivated (approx. 10^5^ gc/mL), and heat inactivated SARS-CoV-2 (approx. 10^5^ gc/mL) suspensions with final concentrations of 50-100 µM photoactivatable dyes (PMAxx™, PEMAX™, or EMA™) or 0·1-2·0 mM platinum compounds (CDDP or PtCl_4_) in DNA LoBind tubes (Eppendorf, Germany). Photoactivation of monoazide dyes was achieved by 10 min of dark-incubation in an orbital shaker (150 rpm) at room temperature (RT) followed by 15 min blue LED light exposure in a photo-activation system (Led-Active Blue, GenIUL). Alternatively, 30 min incubation at RT in an orbital shaker (150 rpm) were used for viability treatments with platinum compounds. A control consisting of genomic RNA or virus suspension without viability marker was included in each assay. Following the viability treatment, the viral RNA was immediately purified as described hereafter.

### Assessment of PtCl_4_ viability RT-qPCR in artificially inoculated and validation in naturally contaminated samples

Platinum (IV) chloride was selected as the most reliable viability marker and tested at final concentrations of 0·5 to 5·0 mM for viability RT-qPCR optimization in stool, urine, nasopharyngeal swabs and wastewater samples.

For the initial optimization, stool and urine specimens that had tested negative for SARS-CoV-2 were retrieved from IATA biobank. Faecal material was resuspended 1% w/v in phosphate-buffered saline (PBS), and supernatant recovered by centrifugation at 2000 × *g* for 5 min. Direct and ten-fold diluted urine, and ten-fold diluted faecal suspension (final 1% *w*/*v* faecal dilution) were spiked with either gamma- and/or heat inactivated SARS-CoV-2 to approximately 10^5^ gc/L final concentration.

Then, nasopharyngeal swabs from positive COVID-19 patients and naturally contaminated wastewater samples were used to validate the viability PtCl_4_ RT-qPCR. Nasopharyngeal swabs (n=9) from COVID-19 positive patients were originally collected at Hospital Clínico Universitario de Valencia (Valencia, Spain) and included in this study once de-identified. To test whether the detection of viral RNA was exclusive for infectious particles, nasopharyngeal swab subsamples were inactivated at 95 °C for 10 min, included in the experiments along with naïve specimen and both assayed by RT-qPCR and viability PtCl_4_ RT-qPCR.

SARS-CoV-2 positive wastewater grabbed samples (n=6) were collected in June-October, 2020 from different wastewater treatments plants involved in a WBE monitoring programme. The samples were originally concentrated by an aluminium precipitation procedure and tested positive for at least two RT-qPCR targets (N1, IP4 or E gene).^(14)^ To exclude additional viral inactivation due to the concentration procedure, wastewater were freshly concentrated by Centricon-Plus 70 centrifugal ultrafilters units with a cut-off of 100 kDa (Merk-Millipore, MA, US).^(15)^ Samples were all diluted in PBS as specified. Viability treatment, RNA extraction and SARS-CoV-2 detection were carried out as hereafter detailed.

### Viral RNA purification and SARS-CoV-2 detection

Viral RNA was extracted using Maxwell® RSC 16 instrument and Maxwell RSC Pure Food GMO and authentication kit (Promega, Spain) and detected by RT-qPCR targeting N1, N2, E gene, IP2 and IP4 regions.^(16)^ Viral RNA from nasopharingeal samples was extracted using a KingFisher™ Flex (Thermo Fisher Scientific) instead. Given the superior sensitivity of N1 RT-qPCR resulting from the initial screening, this target was used for subsequent determinations. Each RT-qPCR assay was performed in duplicate and included nuclease-free water as negative control, and SARS-CoV-2 complete genomic RNA (VR-1986D™, ATCC, VA, US), E gene plasmid (10006896, 2019-nCoV_E Positive Control from Charité/Berlin, IDT, Belgium) or N1/N2 plasmid (10006625, 2019-nCoV_N_Positive Control from CDC, IDT, Belgium) as positive controls. Ten-fold RNA dilutions were consistently tested to check RT-qPCR inhibition due to viability marker residues or inhibitory substances in the sample.

### Statistical analysis

All data were compiled from three independent experiments with at least two technical replicates for each variable. Data are presented as median ± SD. Significant differences in median cycle threshold (Ct) were determined by using either one- or two way(s) ANOVA followed by Dunnett’s multiple comparisons test on GraphPad Prism version 8·02 (GraphPad Software, US). Differences in means were considered significant when the p was <0·05.

## Results

### Initial assessment of viability markers and RT-qPCR assays

With regard to the viability markers tested, platinum compounds were better at preventing PCR amplification of SARS-CoV-2 genomic RNA suspension than monoazide dyes, regardless of the RT-qPCR target (Figure 1). Compared to untreated RNA, significant differences were detected for PtCl_4_ and CDDP treated samples in all the five RT-qPCR targets tested. While PtCl_4_ completely prevented the RNA amplification for all replicates, it occurred in 5 out of 20 CDDP treated replicates targeting N1, N2 and IP4. Among photoactivatable dyes, 50 µM PMAxx offered the best performance as it removed the signal in 8 replicates showing statistically significant differences for E gene, IP2 and IP4. An additional assay tested 100 µM PMAxx on SARS-CoV-2 genomic RNA without any improvement of the results with respect to 50 µM PMAxx concentration (data not shown). EMA and PEMAx completely removed RT-qPCR signals in 2 out of 20 replicates. Given these preliminary results, we further assessed PMAxx and PtCl_4_ effect on SARS-CoV-2 gamma-(ca. 8·50 × 10^5^ gc/mL corresponding to 140 TCID_50_/mL) and heat inactivated (ca. 1·88 × 10^5^ gc/mL corresponding to 80 TCID_50_/mL) viral particles by using N1 as the most sensitive RT-qPCR assay among all the compared targets. PMAxx at 50 µM minimally reduced the PCR signals by 2·82 and 3·17 Cts with respect to the gamma- and heat inactivated controls, while the superior ability of PtCl_4_ was confirmed for both gamma- and heat inactivated SARS-CoV-2 viral particles (Figure 2). A final concentration of 1·0 mM PtCl_4_ was needed to consistently prevent the amplification of inactivated viruses by viability RT-qPCR. Thus, we further applied the PtCl_4_ viability RT-qPCR to high concentrated gamma-inactivated viral suspensions (ca. 8·50 × 10^6^ gc/mL). Results showed that 0·5 and 1·0 mM PtCl_4_ reduced by 3·4 and 6·8 Cts compared to the control. Although significant statistical differences were detected for all treatments regardless of the concentration of the metal compound, only 2·0 mM PtCl_4_ showed to consistently prevent signal amplification (only one positive out of 8 replicates, Ct=39·11).

**Figure 1.**
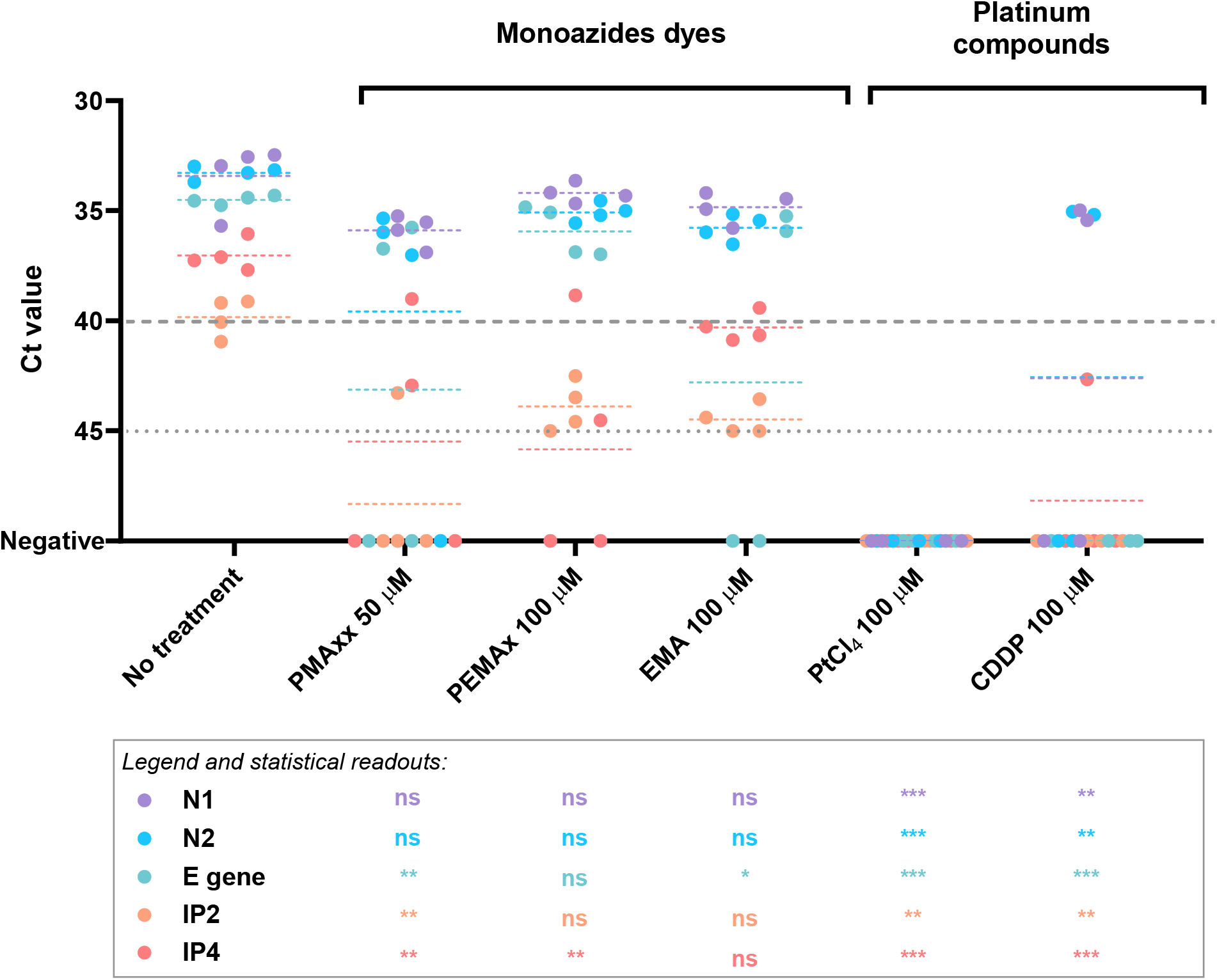
Performance of monoazide photoactivatable dyes and platinum compounds on SARS-CoV-2 genomic RNA assessed by targeting five different RNA regions. Dashed grey line represents RT-qPCR theoretical limit of detection for N1, N2 and gen E; dotted grey line represents RT-qPCR theoretical limit of detection for IP2 and IP4. Asterisks indicate significant difference from untreated control for each molecular target: ns, not significant; *p<0·01; **p<0·001; ***p<0·0001; ns, not significant.

**Figure 2.**
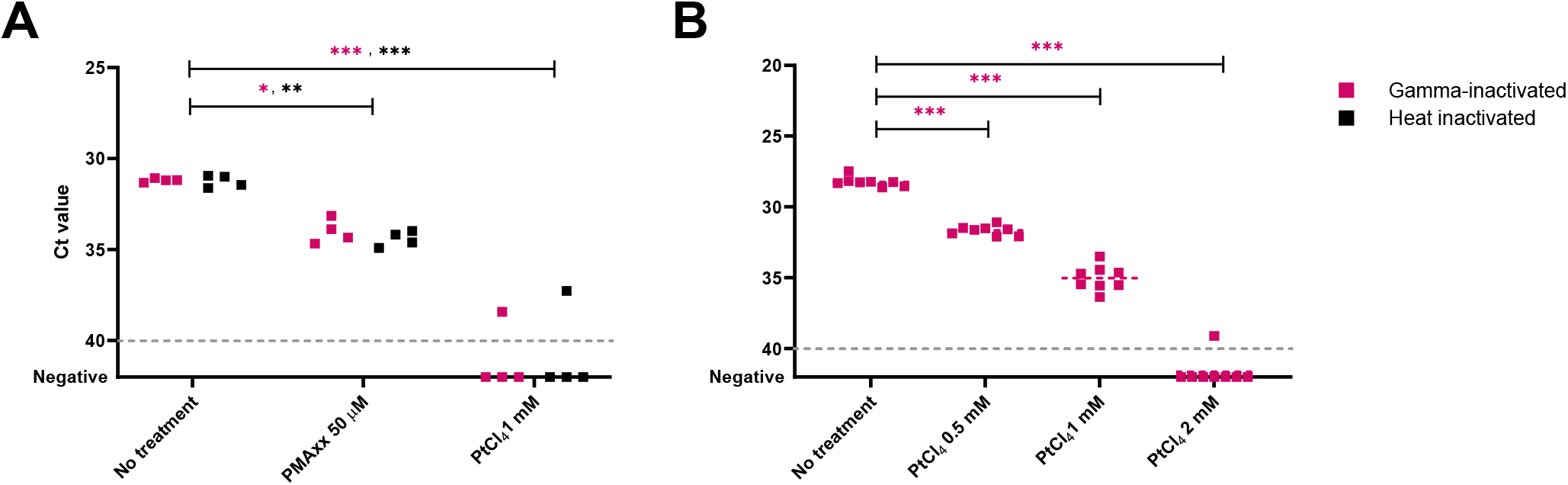
Assessment of viability markers on inactivated SARS-CoV-2 viral particles suspended in PBS buffer at different concentrations. RT-qPCR assays targeted N1 region. A) Comparison of PMAxx and platinum chloride (PtCl_4_) viability RT-qPCRs on low (ca. 10^3^ gc/mL) gamma- and heat inactivated SARS-CoV-2 viral particles. B) Viability RT-qPCR optimization using increasing concentration of PtCl_4_ on high concentrations of gamma-inactivated SARS-CoV-2 viral particles (ca. 10^5^ gc/mL). Dashed grey lines represent RT-qPCR theoretical limit of detection. Asterisks indicate significant difference from untreated control: *p<0·01; **p<0·001; ***p<0·0001.

### Effect of sample complexity on viability RT-qPCR

To determine the effect of sample matrix on viability RT-qPCR, we spiked 10-fold diluted stool suspensions (1% *w*/*v* final dilution) and urine specimen (10% v/*v* final dilution) with approximately 10^5^ gc/mL gamma-inactivated SARS-CoV-2, and applied up to 5·0 mM PtCl_4_ as viability marker. Compared to the untreated control, significant differences were observed for 1·0 mM PtCl_4_ in urine samples or 1·25 mM PtCl_4_ in stool suspensions (Figure 3). However, a concentration of 5·0 mM was needed to completely remove the PCR signals in urine, while 2·5 mM PtCl_4_ prevented the amplification of 1 out of 8 replicates in stool. Although the complete inhibition of amplification signals was achieved to a limited extent, a sharp difference above one logarithm of genomic copies (ΔCts≈3·3) was observed in stool and urine samples processed with 1·25 and 3·75 mM PtCl_4_, respectively.

**Figure 3.**
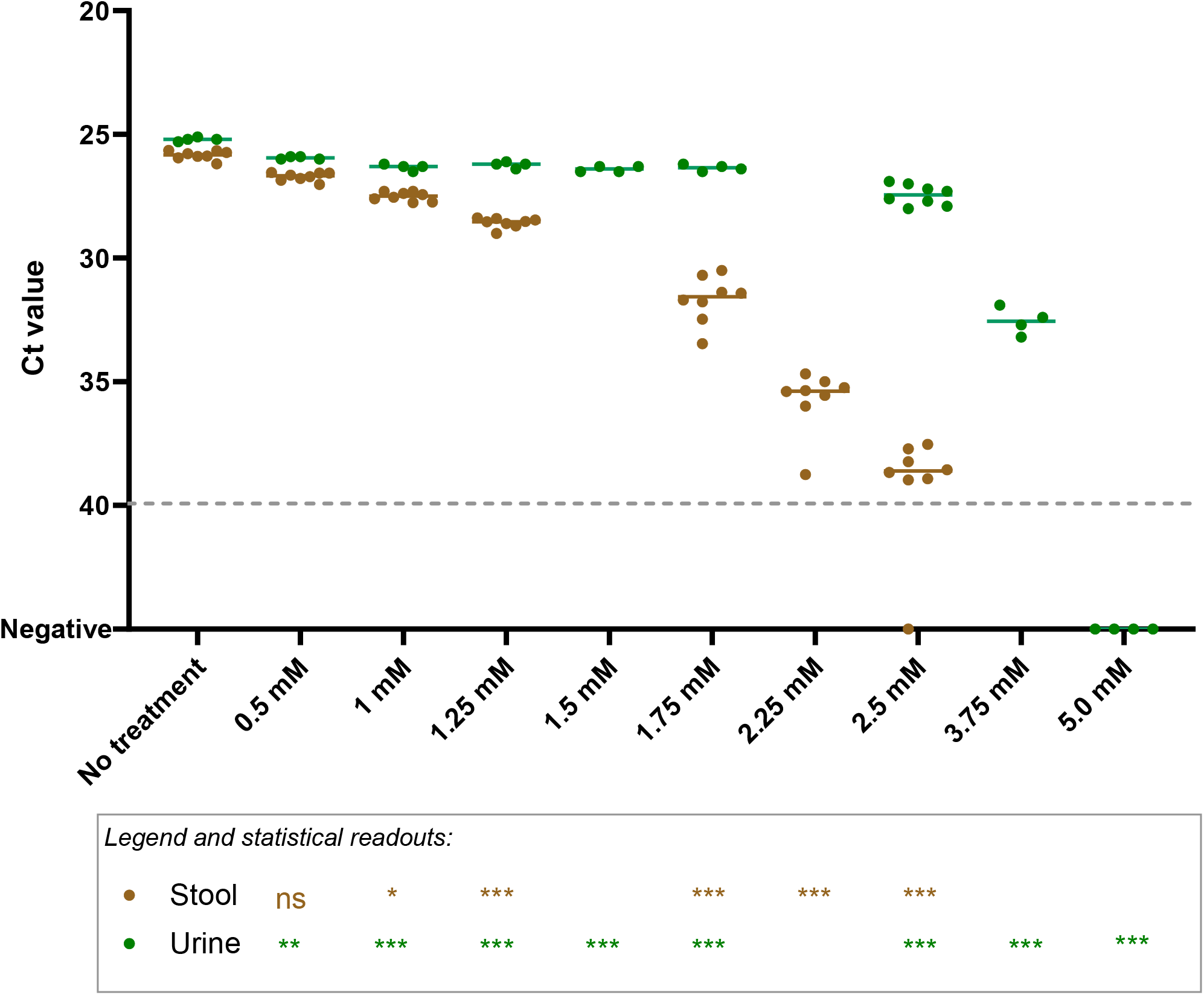
Platinum chloride (PtCl_4_) viability RT-qPCR on ten-fold diluted faecal suspensions (1% w/v final dilution) (brown dots) and urine specimens (10% v/v final dilution) (green dots) spiked with approximately 10^5^ gc/mL gamma-inactivated SARS-CoV-2. Dashed grey line represents RT-qPCR theoretical limit of detection. Asterisks indicate significant difference from untreated control: *p<0·01; **p<0·001; ***p<0·0001; ns, not significant.

### Viability RT-qPCR validation on positive clinical samples and naturally contaminated wastewater

Additional experiments were set up to validate viability PtCl_4_ RT-qPCR on nasopharyngeal swabs from COVID-19 positive patients and on naturally contaminated wastewater samples. Initial experiments using undiluted samples achieved unsuccessful results (data not showed), thus both clinical and wastewater samples were ten-fold diluted in PBS buffer. Nine 10-fold diluted nasopharyngeal swabs and the corresponding heat-inactivated (95 °C for 10 min) subsamples were processed by RT-qPCR alone and viability RT-qPCR with either 1·0 or 2·5 mM PtCl_4_ (Figure 4). Applying 1·0 mM PtCl_4_ viability RT-qPCR, consistent amplification signals were observed in both naïve and heat-treated samples with minimal Ct differences compared to RT-qPCR alone. Increased concentration to 2·5 mM led to a sharper discrimination of PCR signals (ΔCt=9·24 ± 3·59). Similarly, the complete prevention of RT-qPCR signals occurred in one out of four samples at 1·0 mM PtCl_4_, and in three out of five samples at 2·5 mM (Figure 4). Regardless of the viability marker concentration applied, the complete prevention of amplification was observed in samples with initial low viral titer (Ct values ≥ 30). Moreover, the 2·5 mM PtCl_4_ viability RT-qPCR was further validated on six wastewater samples naturally contaminated with SARS-CoV-2. The results showed that 2·5 mM PtCl_4_ completely prevented the amplification in all samples (Figure 5).

**Figure 4.**
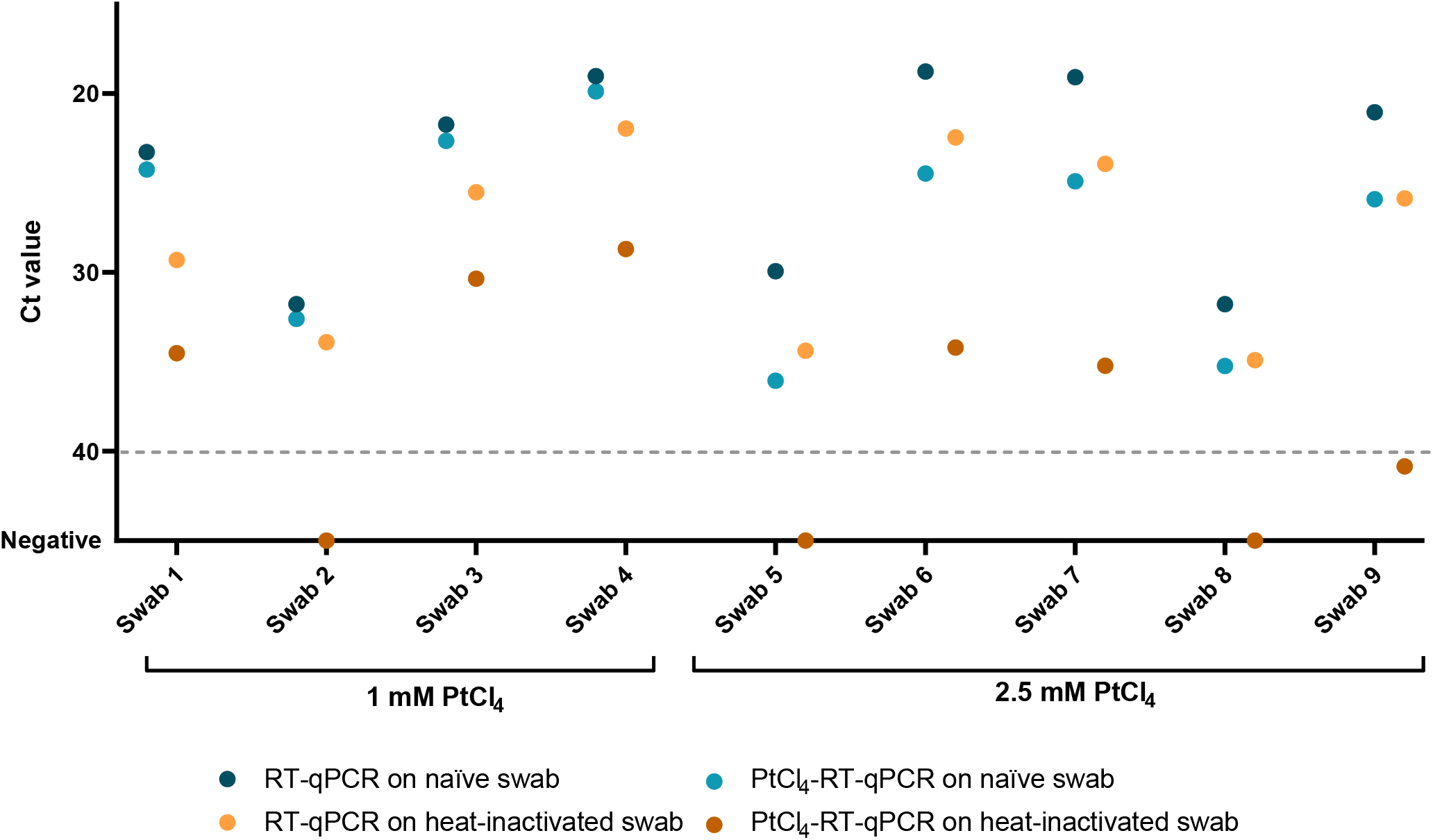
Validation of viability RT-qPCR with either 1mM or 2·5 mM PtCl_4_ on ten-fold diluted nasopharyngeal swabs from COVID-19 positive patients. Plotted dots represents the median cycle threshold value (Ct) of naïve and heat-inactivated (95 °C for 10 min) subsamples assayed by RT-qPCR alone and viability RT-qPCR both targeting N1. Dashed grey line represents RT-qPCR theoretical limit of detection.

**Figure 5.**
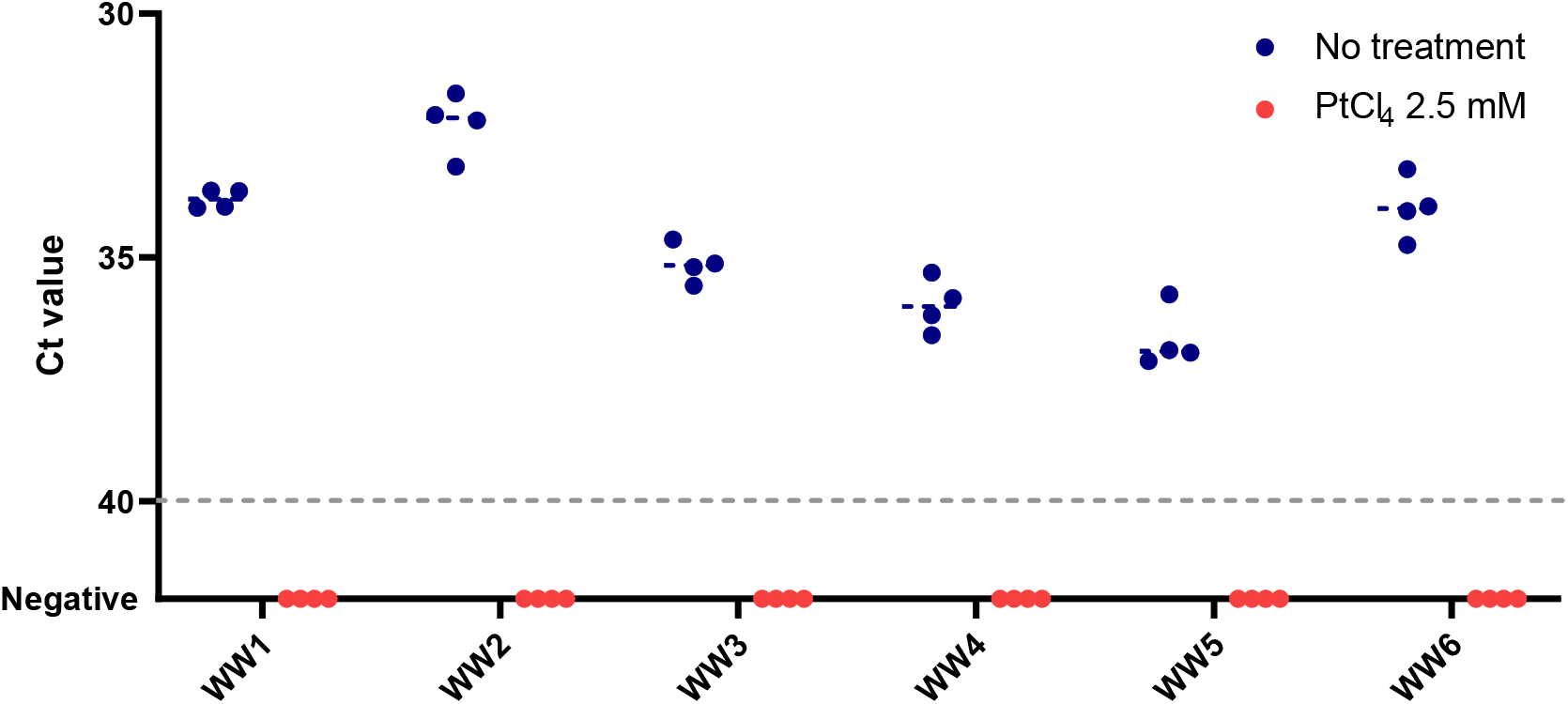
Validation of 2·5 mM PtCl_4_ viability RT-qPCR on ten-fold diluted naturally contaminated wastewater samples. Dashed grey line represents RT-qPCR theoretical limit of detection.

## Discussion

Currently, research projects aiming to assess the risk of transmission and exposure to infectious virus either in clinical and environmental settings have been limited by the biosafety level-3 (BSL-3) conditions needed to handle infectious SARS-CoV-2. The research effort of this investigation intended to provide a rapid and sensitive analytical method that selectively detects potentially infectious SARS-CoV-2 in a significantly shorter time than the traditional cell-culture based method and that can be used in a wide range of applications, including clinical and environmental COVID-19 monitoring programs as epidemiological response to the pandemic that is causing such a public health emergency.

This study evaluated photoactivatable monoazide dyes and metal compounds as viability markers applied prior to nucleic acid extraction to prevent amplification of RNA from non-viable viral particles, thus enabling amplification only of viable/infectious viruses in downstream RT-qPCR assay. Selecting platinum chloride as the best performing viability marker, we demonstrated that viability RT-qPCR efficiently discriminated free RNAs and inactivated SARS-CoV-2 inoculated in buffer, stool and urine suspensions. Then, we further proved that the method inferred SARS-CoV-2 infectivity better than RT-qPCR alone in both nasopharyngeal swabs from positive COVID-19 patients and in naturally contaminated wastewater samples. In the case of complex matrices, increased PtCl_4_ concentration of 2·5 mM and ten-fold sample dilution are recommended because of the presence of suspended solids and inhibitors that hinder the efficacy of the viability treatment.

Our investigation initially included five well-established molecular assays since the length of the amplicon and/or the richness of secondary structures of targeted RNA may affect the efficiency of viability treatments.^(17,18)^ We show that metal compounds performed better than monoazide dyes, irrespective of RT-qPCR assays. However, RT-qPCR targeting N1 region is recommended because of its superior sensitiveness. This aspect is of importance because complex matrices needed to be diluted to achieve a more efficient inference of viral infectivity, as demonstrated in spiked stool and urine, in positive swabs and in contaminated wastewater. Similarly, sample dilution was needed to implement a viability RT-qPCR targeting norovirus in sewage.^(19)^ Moreover, N1 assay better fits the testing on samples with expected low viral concentrations (e.g., environmental samples) and/or PCR inhibitors (e.g., concentrated wastewater, stool). As N1 assay has been validated in many laboratories worldwide, this viability method could also be easily and widely implemented.

To the best of our knowledge, this is the first report on a rapid molecular assay independent from viral replication in cell culture developed to test SARS-CoV-2 infectivity. A recent investigation by our group demonstrated the suitability of viability RT-qPCRs to infer the infectivity of porcine epidemic diarrhea coronavirus, member of the genus *Alphacoronavirus* within *Coronaviridae* family.^(13)^ Interestingly, we were able to demonstrate that PMAxx viability RT-qPCR matched the thermal inactivation pattern obtained by cell culture better than other viability markers, including PtCl_4_, and RT-qPCR alone.

Alternative rapid methods to assess the viability of enveloped viruses have been explored with unsuccessful results. These included propidium monoazide and immunomagnetic separation tested on laryngotracheitis virus and ethidium monoazide on avian influenza virus.^(11,12)^ It is worth to report that we also tested on inactivated SARS-CoV-2 suspensions a porcine gastric mucine *in situ* capture RT-qPCR, a method that was originally implemented in our laboratory for human enteric viruses.^(20)^ Although proper SARS-CoV-2 infectious controls could not be included, those experiments resulted in inconclusive outcomes (data not shown).

Our viability RT-qPCR results of nasopharyngeal swabs from positive COVID-19 patients indicate the potential infectivity of the samples, while naturally contaminated wastewater are unlikely to contain infectious viral particles. These later findings reflect the viral replication in cell culture from RNA positive stool and respiratory samples as well as the unsuccessful attempts to isolate and cultivate SARS-CoV-2 from wastewater samples.^(21–23)^

Our findings are clinically relevant as RT-qPCR has become the primary method to diagnose COVID-19. However, as it detects RNA, its ability to determine the infectivity of patients is limited.^(5–8)^ In addition, the immune system can neutralise SARS-CoV-2 preventing subsequent infection but not eliminating nucleic acid, which degrades slowly over time. This has been confirmed in cohort studies that concluded that seroconversion does not necessarily lead to the elimination of viral RNA, with cases being RT-PCR positive up to > 63 days after symptom onset despite having neutralizing antibodies.^(24–26)^ Furthermore, some reports correlated the infectiousness of upper respiratory tract samples with RT-qPCR Ct values in COVID-19 cases. Analysing a large dataset (n=324), Singanayagam and colleagues demonstrated that the probability of viral recovery from samples with 27.5 < Ct < 30 was ≈66%, decreasing to ≈ 28% for 30 < Ct < 35, and to 8·3% for Ct > 35.^(23)^ Similarly, Bullard observed SARS-CoV-2 cell infectivity only for respiratory specimens sampled < 8 days symptom onset with Ct < 24.^(27)^ However, viral replication was also obtained from samples with elevated Ct values of 36-39.^(7,28)^ Notwithstanding, the correlation between RNA and virus isolation remains unclear.

Unfortunately, we had no access to clinical samples with higher Ct values which are likely to contain non-infectious particles to contrast such hypothesis by the proposed viability RT-qPCR. In addition, the ratio between viral shedding and infectivity has been reported to vary along the course of the infection.^(4,6,22)^ This information regarding epidemiological characteristics, symptom history and relevant sampling details included in medical records could have explained at least in part the different performances of viability RT-qPCR among clinical samples, however it could not be retrieved as de-identified specimens were analysed in this study.

Despite the viability treatment, we detected residual signals in heat inactivated nasopharyngeal swabs. This could be attributed to the viral envelop and nucleoproteins that limit the access and/or the binding of viability markers to SARS-CoV-2 RNA, as hypothesised for avian influenza virus and bacteriophage T4. ^(12,29)^ The enveloped structure of coronaviruses may also explain the increased concentration of viability markers needed for SARS-CoV-2 and PEDV compared to human enteric viruses.^(13,19,30)^ This cumbersome finding obtained by the proposed viability procedure suggests that the overestimation of the infectivity of a given sample may occur which, although warranting a careful interpretation, represents a conservative prediction. With regards to wastewater samples that tested positive for SARS-CoV-2 RNA, they most probably contained detergents and chemicals that are detrimental to viral infectivity further supporting the efficacy of the viability RT-qPCR in discriminating potentially infectious and inactivated viral particles.^(31)^ The ultimate confirmation on the infectivity of the samples by cell culture, although recommendable, could not be provided. Detection of SARS-CoV-2 by either culture and viability RT-qPCR is valuable as a proxy for infectiousness; however, as the human infectious dose remains unknown, the significance of low titres of infectious virus for human-to-human transmission remains uncertain. Above all, as some individuals reportedly remain PCR positive weeks after SARS-CoV-2 infection recovery, knowing whether viral RNA in these persistent carriers is contagious provides key insights for quarantine policy, to safely discontinue self-isolation and contact tracing as essential public health measures to definitively prevent transmission.^(6,26,32)^ Besides some limitations, the proposed viability RT-qPCR effectively reduced the amplification signals of non-infectious and free RNA of SARS-CoV-2 in complex matrices finally providing a better estimation of the infectiousness of samples. Thus, mathematical models derived from laboratory scale experiments comparing viability RT-qPCR and viral replication could correlate viral load and infectivity, finally providing relevant tools of interest based on rapid molecular assay for prevention strategies and risk assessment.

## Conclusions

In conclusion, the use of pre-treatments to prevent RT-qPCR amplification of RNA from non-infectious SARS-CoV-2 using platinum chloride as a viability marker of infectivity was implemented in stool and urine samples and successfully validated in naturally contaminated wastewater samples, supporting the idea that SARS-CoV-2 present in sewage is not infectious. Residual amplification signals in nasopharyngeal swabs exposed to heat-inactivation overestimated the amount of viable virus, still providing a conservative interpretation of the infectiousness of the sample. Our study proposes a rapid analytical tool based on viability RT-qPCR to infer SARS-CoV-2 infectivity with potential application in risk assessment, and prevention and control in public health programmes.

## Contributors

GS, AA and WR conceived the study with input from AP-C, IF, SML and EC-F. EC-F, IF, AD-R, and IG-G led data acquisition. DN provided the nasopharyngeal swabs from COVID-19 positive patients. GS, EC-F, and WR led data analysis, interpretation, and visualisation. EC-F and WR wrote the draft report, and all other authors revised the report critically for important intellectual content. All authors critically reviewed and approved the final version of the manuscript.

## Data Availability

Data available under request

## Acknowledgements

The study was supported by CSIC (202070E101), Generalitat Valenciana (Covid_19-SCI), MICINN co-founded by AEI/FEDER, UE (AGL2017-82909), and MICINN/AEI (PID2019-105509RJ-I00). EC-F is recipient of a predoctoral contract from the MICINN, Call 2018.

We thank Agustin Garrido Fernández and Andrea Lopez de Mota at IATA-CSIC for providing support in sample processing. We acknowledge Global Omnium S.L., NILSA, Aguas de Malaga, ESAMUR and FACSA for coordinating and managing wastewater sampling.

## Research in context

### Evidence before this study

Previous works have established RT-qPCR methods for the efficient detection of severe acute respiratory syndrome coronavirus 2 in stool, urine and respiratory samples, with the latter being the most commonly used specimens in tracing COVID-19 cases. Similarly, wastewater-based epidemiology has been globally implemented to monitor the community spread of the virus. However, SARS-CoV-2 RNA load in clinical and environmental samples is not indicative of infectiousness. The distinction between viral shedding and infectivity is important for the development of prevention policy and quarantine guidelines as well as for the assessment of the risk of contamination in environmental settings. Some reports correlated RT-PCR Ct value and culture positivity and there is scientific evidence that persistent PCR positive individuals are not transmitting at the post-symptomatic stage of infection. However, testing viral replication is not feasible for large scale clinical screening or for environmental surveillance. In addition, the biosafety level 3 laboratory requirements needed to handle SARS-CoV-2 infectious materials and the turnaround time for culture results has also limited investigation on related topics. Previous works have established rapid viability assays based on RT-qPCR pre-treatments to infer the potential infectivity of enteric viruses, including norovirus, hepatitis A and E viruses in environmental and food samples. Unsuccessful attempts were reported for influenza virus and infectious laryngotracheitis virus, while a viability RT-qPCR for the selective detection of infectious and heat-inactivated porcine epidemic diarrhea coronavirus, as an enveloped viral model, was recently proposed. However, the sensitivity and reliability of this method remains to be shown for SARS-CoV-2.

### Added value of this study

We show that SARS-CoV-2 RNA amplification of non-infectious samples can be reproducibly prevented by processing clinical and wastewater samples by a platinum chloride viability RT-qPCR technique. To the best of our knowledge this is the first report on this subject that also provides the following distinguishing features: (i) the procedure was optimized using standard materials which include SARS-CoV-2 genomic RNA, gamma-and heat inactivated viral particles; (ii) the preliminary findings were validated in nasopharyngeal swabs from COVID-19 positive patients and in wastewater samples naturally contaminated with SARS-CoV-2 RNA.

Overall, this study allows to selectively detect SARS-CoV-2 RNA belonging to potentially infectious viral particles in complex matrices of interest for epidemiological surveillance and pandemic control.

### Implications of all the available evidence

Extensive lockdown measures are currently allowing to partially mitigate the spread of SARS-CoV-2. As such, it is of extreme importance to set up feasible and reliable epidemiological surveillance strategies that include clinical and environmental monitoring programs. Information on the infectivity of such samples is the keystone to better define the transmission route(s) and the risk of exposure. Further developments of predictive models based on such a viability approach may have a broad range of applications: including risk assessment, food safety and public health policy.

